# Temporal trends in legionellosis national notification data and the effect of COVID-19, Switzerland, 2000- 2020

**DOI:** 10.1101/2022.01.19.22269395

**Authors:** Fabienne B. Fischer, Daniel Mäusezahl, Monica N. Wymann

**Affiliations:** Swiss Tropical and Public Health Institute, Basel, Switzerland; University of Basel, Basel, Switzerland; Federal Office of Public Health, Berne, Switzerland

**Keywords:** Legionnaires’ Disease, Legionellosis, Switzerland, COVID-19, Disease Surveillance, Communicable Diseases

## Abstract

The notification rate of legionellosis in Switzerland and other European countries has markedly increased over the last 20 years. Here, we investigated the Swiss notification data on legionellosis from 2000-2020 in regards of overall time trend, content and data quality. We further explored the impact of the COVID-19 pandemic on the reported case numbers using an interrupted time series approach. Between 2000 and 2020, 5,980 cases were included in our analysis. The annual crude notification rate for legionellosis cases increased from 1.1/100,000 population (CI: 0.9 - 1.4) in 2000 to 5.6/100,000 population (CI: 5.1 - 6.1) in 2020. In recent years, the summer peaks have been more pronounced and some shifted earlier in the year. The highest notification rate was recorded in 2018 with 6.7/100,000 population (CI: 6.2 - 7.3). The hospitalisation rate for notified cases remained high across all study years (89.9%), while the case fatality rate slightly decreased (from 7.7% to 3.6%). COVID-19 containment measures, such as travel restrictions and/or related behavioural changes, are associated with a temporary decline in cases of 35%. Overall, the quality of the notification data was good. Clinical data were more susceptible to interferences than data from laboratory reporting, which could be observed most clearly in the decline of clinical reports by 4.3 percentage points in 2020. As the case classification for Legionnaires’ disease includes pneumonia symptoms, this decline could lead to an underestimation of Legionnaires’ disease cases, yet the continuous reporting though the diagnostic laboratories suggested a robust surveillance system for legionellosis in Switzerland.

## 1 Introduction

The term legionellosis comprises all diseases caused by *Legionella* spp. The majority of the known burden of disease stems from Legionnaires’ disease (LD), which presents in a form of severe pneumonia. Legionellosis is caused by inhalation or aspiration of aerosols from contaminated water sources, and has the potential to occur as larger outbreaks, even though most cases are sporadic. To detect such outbreaks, monitor disease trends, and take appropriate public health measures, legionellosis is included in the passive disease surveillance system of many, mostly high-income, countries [1].

In the last two decades, the notification rate of legionellosis steadily increased in Switzerland, other European countries and the US [2, 3]. Several hypotheses for the increase in disease incidence were formulated such as changes in weather and climate, changes in energy policy and buildings / water systems infrastructure, both thought to promote *Legionella* spp. growth, and, demographic changes with an increasing susceptible population for LD [2, 4]. Yet, the observed disease trend is not only shaped by changes in incidence, but also prone to react to any changes in the processes leading up to the case being reported, e.g. health-seeking behaviour, diagnosis and reporting procedures [5].

In Switzerland, cases of legionellosis are notifiable to the National Notification System for Infectious Diseases (NNSID) since December 1987. The NNSID is managed by the Federal Office of Public Health (FOPH). Trigger for a mandatory notification is a positive confirmation for a *Legionella* spp. infection. If testing is done by a laboratory, the diagnostic laboratory has to notify simultaneously to the cantonal health authorities and the FOPH with the “report on laboratory findings”. This laboratory report triggers a “report on clinical findings” from the treating physician to the cantonal health authorities. The cantonal health authorities check for completeness of the clinical information provided and if immediate measures are necessary. They then forward the information to the FOPH. If testing is done by rapid urine antigen test (UAT) without involvement of a diagnostic laboratory, the “report on clinical findings” indicates the test result, in the absence of a laboratory report. At the FOPH, the paper-based clinical and laboratory notification forms are recorded electronically and are matched by patient. The timeframe for reporting of both laboratory and clinical findings for legionellosis is one week [6].

Before 2000, there were substantial changes to the notification process, hampering the evaluation of prior disease trends. Since then, there were only few adjustments made to the notification form and to the case classification for LD, which was last updated in 2012 (see Table 1) [7]. Cases classified as “possible” were either without pneumonia or without clinical information on pneumonia. They count towards legionellosis cases, but not as LD. Since 2006, the FOPH also requested diagnostic laboratories to report the annual number of tests performed for *Legionella* spp. to obtain complementary denominator data to improve contextualisation of the surveillance data [7]. The quality of this reporting, however, was insufficient; therefore, a research study investigated the positivity for the years 2007-2016 [8]. The authors found a strong and parallel increase of the test volume and the number of positive cases. However, without an assessment of the reasons for the increase in test volume, a conclusion on the observed notification trend could not be made.

**Table 1.** Case definition for Legionnaires’ disease in Switzerland since 2012 [7].

| Case classification |  |
| --- | --- |
| Confirmed case | Any person meeting the clinical criterion AND at least one laboratory criteria for a confirmed case |
| Probable case | Any person meeting the clinical criterion AND at least one laboratory criteria for a probable case |
| Possible case | Any person meeting at least one of the laboratory criteria for either a confirmed or probable case AND missing information on the clinical criterion OR clinical criterion not met |
| Criteria |  |
| Clinical criterion | Any person with pneumonia |
| Laboratory criteria for a confirmed case | Either isolation of *Legionella* spp. from respiratory secretion or any normally sterile site OR detection of *Legionella pneumophila* antigen in urine |
| Laboratory criteria for a probable case | Detection of *Legionella* spp. nucleic acid in clinical samples (using for example PCR) OR detection of *Legionella pneumophila* antigen for example by DFA staining using monoclonal-antibody-derived reagents OR significant rise in specific antibody level to *Legionella pneumophila* or other *Legionella* spp. in paired serum samples OR single high level of specific antibody to *Legionella pneumophila* serogroup 1 in serum. |

The COVID-19 pandemic in 2020 has affected the notification rates of almost all mandatory notifiable diseases in Switzerland, including LD [9]. LD cases in 2020 reduced by one third (-33%, 95% CI -43 – -19%) compared to the expected number of LD cases based on the five years prior to the pandemic. Multiple mechanism could explain the impact of the pandemic on LD: First, changes in people’s behaviour could affect incidences and health-seeking behaviour; second, the clinical presentation of LD being similar to COVID-19 [10] could lead to higher testing rates and third, the heavy burden on the health care system could affect testing and reporting behaviours. In particular, the ubiquitous travel and entry restrictions were hypothesised to have reduced cases of travel-associated Legionnaires’ disease (TALD) [11]. Additionally, the closure of public buildings for leisure activities e.g. sport centres, shopping malls, office buildings, and schools might have reduced exposure during the closure but could have led to increased proliferation of *Legionella* spp. in the then stagnant water in unused buildings. Upon re-opening and without thorough flushing of the pipes, the risk for an infection is thought to be increased [12-15]. However, as of now, there has been no quantification of this effect.

Additionally, in 2017, thresholds of *Legionella* spp. contamination in potable, publicly accessible water were regulated in the Food Safety Law [16]. Consequently, *Legionella* spp. became a new concern for the Federal Food Safety and Veterinary Office (FSVO). Due to these developments in the past years, the increasing attention towards legionellosis and efforts to understand and prevent illness cases, an analysis of the past 20 years of LD notification in Switzerland is timely. The first aim of this study is to describe the Swiss notification data for LD for the two decades between 2000 and 2020, specifically the content of the notification data (i.e., cases per week and their characteristics), and the quality of the data (i.e., completeness, validity and timeliness). The second aim is to explore the impact of the COVID-19 pandemic on the content and the quality of these data.

## 2 Methods

### 2.1 Study design and setting

This is a cross-sectional retrospective study utilising routinely collected health data for legionellosis from the NNSID in Switzerland between 01.01.2000 and 31.12.2020. The year 2000 was chosen as the starting time point, as there have been significant changes to the notification system earlier on, rendering older data incomparable.

### 2.2 Legionellosis notification data sources, access and processing

The raw data presented by the NNSID [17] reports all legionellosis notifications before case classification, including cases later classified as possible and “no case”, irrespective of their residency. After classification based on the case definition shown in Table 1, the FOPH retains only confirmed and probable cases, i.e., LD cases, with residency in Switzerland or the Principality of Liechtenstein, in their reports. For the purpose of this study, we used the same inclusion criteria for residency, but kept confirmed, probable and possible cases in the dataset and only excluded “no cases”.

The legionellosis notification data underwent the routine cleaning processes at the FOPH. For data confidentiality reasons, variables like date of birth and place of residence are stored in separate files and deleted after three years. For the years 2000 to 2016, we therefore, obtained only the age in years and the canton of residence. We did not exclude case records that violated the internal validity (illustrative example: an observation with the hospitalisation date after the death date), in order to present the full dataset and explore its quality.

The legionellosis notification dataset contained cases notified on any given day. Due to low case numbers and to eliminate the effect of the day of the week on health-seeking behaviour and case confirmation, we aggregated data on a weekly level. The case notification further contains information on the patient’s demographics (date of birth, sex, residential address, nationality), clinical information (date of disease onset and diagnosis, hospitalisation status, death), diagnostic information (sample material and diagnostic method), information about exposures prior to disease onset (locations, activities, installations), risk factors for development of LD, and information about the notification process (date of data entry, case classification, number of received notification forms).

Age categories were pre-set by the FOPH according to the standard of the European Center for Disease Prevention and Control (ECDC). The FOPH further categorises cases based on the most probable exposure in the 2-10 days prior to onset of illness: travel-associated, retirement-home-associated, nosocomial, professional-associated, and community-acquired [18]. Community-acquired cases include both, cases with a probable or confirmed infection in the community and cases, without another exposure category indicated.

### 2.3 Quantification of the impact of the COVID-19 pandemic on legionellosis cases

To address the second aim, the exploration of the impact of the pandemic, we collected information on the development of the COVID-19 pandemic, either quantitative (case numbers, hospitalisations, deaths and tests) or qualitative (non-pharmaceutical interventions implemented).

Information on the evolution of the COVID-19 pandemic in Switzerland were taken from the Oxford COVID-19 Government Response Tracker (OxCGRT) [19], which has been adapted for the Swiss context [20]. This information was complemented with our own compilation of events. Data on the number of COVID-19 cases, hospitalisation, deaths and testing is publicly available and was extracted on 4 February 2021 [21]. Data on the COVID-19 pandemic contains daily information from the start of the pandemic in Switzerland (early February 2020) until end of December 2020 and was also aggregated by week.

### 2.4 Linkage of legionellosis case data and COVID-19 data

For the legionellosis data, to identify events and cases on the timeline, we used the variable “case date”, which is generated within the NNSID. The case date denotes the earliest date available from a series of date-related variables per case. Ideally, and in most cases, this is the date of symptom onset. The OxCGRT and COVID-19 case database had unique time identifiers, which allowed linkage with the LD database on the timeline.

We used population statistics from the Swiss Federal Statistical Office (FSO) to calculate crude and adjusted notification rates. At the time of the analysis, these statistics were not yet available for 2020; therefore, we used the statistic from 2019 instead.

### 2.5 Statistical methods

#### 2.5.1 Descriptive analyses

Data were descriptively analysed in terms of data content and data quality using the statistical software R (Version 1.3.1093 [22]). Notification rates, defined as the number of notified cases per 100′000 resident population, were calculated using population statistics from the FSO. Confidence intervals for crude rates were calculated using the package propCIs using exactci to apply the Clopper-Pearson exact CI approach. Confidence intervals for adjusted rates have been calculated using the package dsrTest to apply the Gamma Method proposed by Fay and Feuer (1997) [23].

#### 2.5.2 Interrupted time series analysis

To address the second aim, we used an interrupted time-series analysis (ITSA) approach as outlined by Bernal, Cummins and Gasparrini (2017) to estimate the effect of selected measures on the legionellosis case numbers [24]. The selected events were i) the implementation and lifting of travel restrictions, on 16^th^ March (week 12) and 15^th^ June (week 25), and ii) the opening of schools and leisure activity facilities on 11^th^ of May (week 20) after almost two months of closure [25]. As there has been stepwise openings, we excluded the data points during the opening phase from week 20 until week 24. We hypothesised a lagged step-change for both events. With count data available, we modelled the weekly number of cases between 2016 and 2020 using a quasi-Poisson regression model with the standardised population as the offset. We incorporated harmonic functions to account for seasonality and a lag-time of one week (incubation time) into the model [26].

## 3 Results

### 3.1 Time trend in legionellosis cases

Figure 1 shows the increasing weekly case numbers since 2000 until 2018, followed by a small drop in 2019 and 2020. The annual crude notification rate for legionellosis cases ranged from 1.1/100,000 population (CI: 0.9 - 1.4) in 2000 to 5.6/100,000 population (CI: 5.1 - 6.1) in 2020. The highest notification rate was recorded in 2018 with 6.7/100,000 population (CI: 6.2 – 7.3).^1^

**Figure 1.**
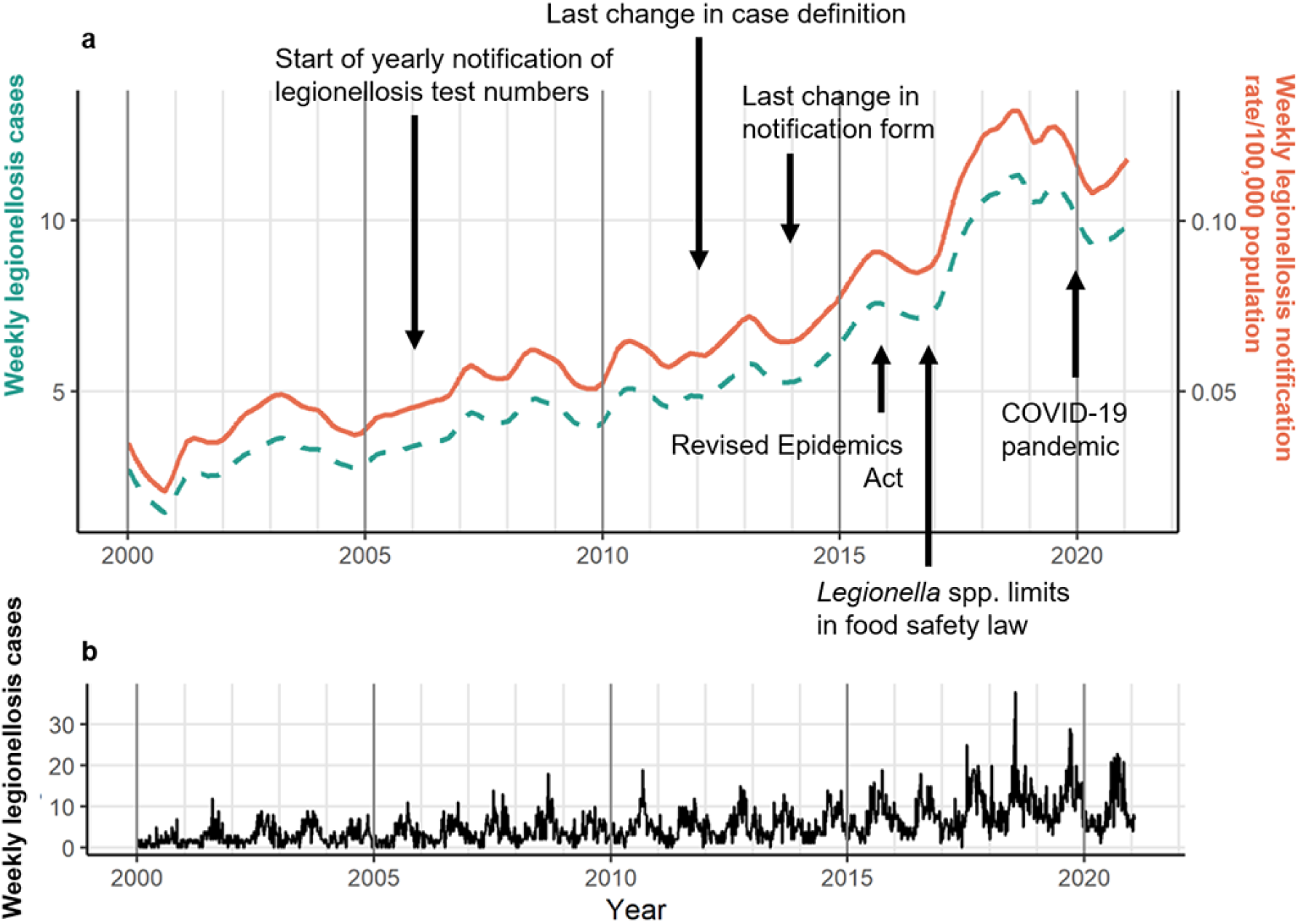
**a** Time trend (without seasonality and randomness) of legionellosis cases in Switzerland, 2000-2020. **b** Complete times series of legionellosis cases including trend, seasonality and randomness.

There is a strong annual seasonality in the data peaking around calendar week 36 (Figure 2). The record-high year of 2018 showed a strong summer peak, which however, shifted to June instead of August. Since 2000, the increase of cases in the summer months has been more pronounced than the increase during the winter months. Comparing the period 2010-2015 with the period 2016-2020, the number of cases increased most strongly in spring (Mar - May) by 85.1%. The cases during summer (Jun - Aug) increased by 75.3%, compared to an increase of 53.3% during autumn (Sept - Nov) and 58.7% during winter (Dec –Feb).

**Figure 2.**
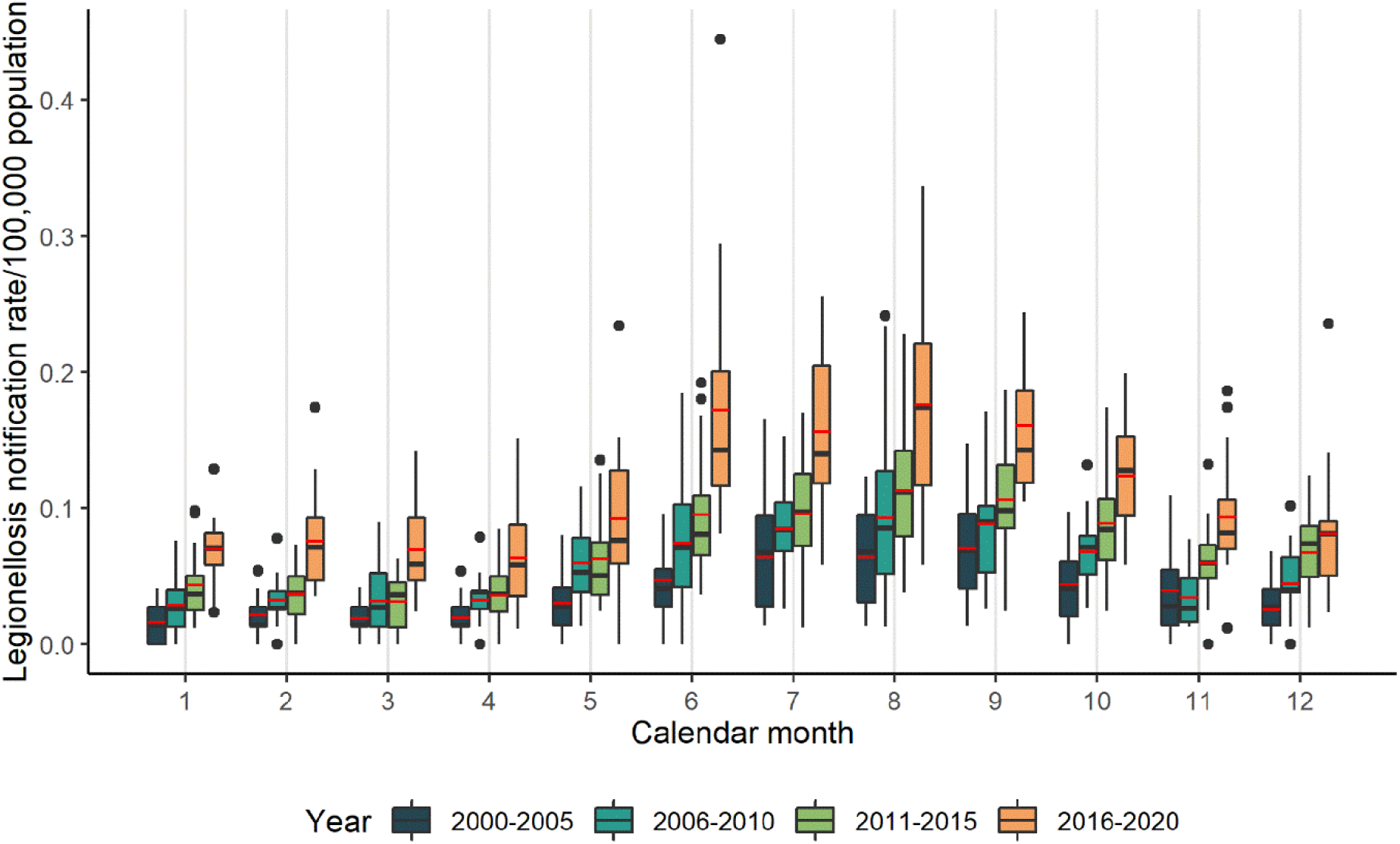
Seasonality of legionellosis cases in Switzerland, 2000-2020. The red line in the boxplot denotes the mean, the black line the median. The black dots denote outliers.

### 3.2 Content of notification

#### 3.2.1 Demographics

Between 2000 and 2020, the database of the NNSID included 5,980 legionellosis cases. Table 2 shows a comparison of the key variables across the years.

**Table 2.** Key variables across the years for notification of legionellosis in Switzerland, 2000-2020

|  | **2000-2005** |  | **2006-2010** |  | **2011-2015** |  | **2016-2020** |  | **Overall** |  |
| --- | --- | --- | --- | --- | --- | --- | --- | --- | --- | --- |
|  | [%] | *N* | [%] | *N* | [%] | *N* | [%] | *N* | [%] | *N* |
| ***Notification*** |  |  |  |  |  |  |  |  |  |  |
| Confirmed case of LD | 89.0 | 784 | 87.0 | 978 | 90.9 | 1,353 | 92.1 | 2,288 | 90.4 | 5,404 |
| Probable case of LD | 4.5 | 40 | 3.9 | 44 | 4.3 | 64 | 1.0 | 26 | 2.9 | 174 |
| Possible case of LD | 6.5 | 57 | 9.1 | 102 | 4.8 | 72 | 6.9 | 171 | 6.7 | 402 |
| Clinical criteria for case definition fulfilled | 93.6 | 825 | 91.7 | 1,031 | 95.5 | 1,421 | 93.6 | 2,325 | 93.7 | 5,603 |
| Laboratory criteria fulfilled | 100.0 | 881 | 99.8 | 1,122 | 99.7 | 1,484 | 99.1 | 2,462 | 99.5 | 5,950 |
| ***Demographics*** |  |  |  |  |  |  |  |  |  |  |
| Median age and 95% CI | 63 | (61-64) | 63 | (62-64) | 64 | (63-65) | 65 | (64-66) | 64 | (63-64) |
| Female | 32.5 | 286 | 29.4 | 330 | 30.4 | 452 | 31.8 | 791 | 31.1 | 1,858 |
| Swiss nationality | 72.2 | 636 | 65.2 | 733 | 71.8 | 1,068 | 69.7 | 1,733 | 69.8 | 4,171 |
| ***Seasonality*** |  |  |  |  |  |  |  |  |  |  |
| Spring (Mar, Apr, May) | 14.2 | 125 | 18.4 | 207 | 14.9 | 222 | 16.6 | 411 | 16.1 | 965 |
| Summer (Jun, Jul, Aug) | 37.6 | 331 | 37.2 | 419 | 35.7 | 531 | 37.5 | 931 | 37.0 | 2,212 |
| Autumn (Sep, Oct, Nov) | 34.5 | 304 | 28.8 | 324 | 31.2 | 465 | 28.7 | 713 | 30.2 | 1,806 |
| Winter (Jan, Feb, Dec) | 13.7 | 121 | 15.6 | 174 | 18.1 | 270 | 17.2 | 428 | 16.6 | 993 |
| ***Region*** |  |  |  |  |  |  |  |  |  |  |
| Central Switzerland | 4.2 | 37 | 6.3 | 71 | 6.8 | 101 | 7.6 | 189 | 6.7 | 398 |
| Eastern Switzerland | 8.4 | 74 | 6.9 | 78 | 11.4 | 169 | 9.5 | 237 | 9.3 | 557 |
| Espace Mittelland | 22.2 | 196 | 20.8 | 234 | 22.3 | 332 | 20.4 | 506 | 21.2 | 1,268 |
| Lake Geneva | 20.3 | 179 | 19.7 | 221 | 21.6 | 321 | 18.5 | 459 | 19.7 | 1,180 |
| Northwestern Switzerland | 14.6 | 129 | 16.3 | 183 | 11.1 | 165 | 13.9 | 346 | 13.8 | 823 |
| Ticino | 14.9 | 131 | 15.7 | 176 | 13.9 | 207 | 15.3 | 379 | 14.9 | 894 |
| Zurich | 15.0 | 132 | 14.0 | 157 | 12.6 | 188 | 14.5 | 360 | 14.0 | 838 |
| ***Clinic*** |  |  |  |  |  |  |  |  |  |  |
| Hospitalisations | 86.5 | 762 | 86.9 | 977 | 89.3 | 1,329 | 84.7 | 2,104 | 86.5 | 5,173 |
| Deaths | 6.4 | 56 | 6.9 | 78 | 4.4 | 66 | 4.0 | 100 | 5.0 | 300 |
| **Exposition** |  |  |  |  |  |  |  |  |  |  |
| Old-age home | 2.7 | 24 | 2.8 | 32 | 3.3 | 49 | 2.6 | 64 | 2.8 | 169 |
| Community-acquired | 72.5 | 639 | 80.9 | 909 | 76.9 | 1,144 | 80.0 | 1,987 | 78.3 | 4,681 |
| Nosocomial | 6.0 | 53 | 3.8 | 43 | 3.5 | 52 | 3.4 | 84 | 3.9 | 231 |
| Occupational | 1.7 | 15 | 1.4 | 16 | 1.6 | 24 | 2.1 | 51 | 1.8 | 106 |
| Travel-associated | 17.0 | 150 | 11.0 | 124 | 14.7 | 219 | 12.0 | 299 | 13.3 | 793 |
| **Risk factors for LD** |  |  |  |  |  |  |  |  |  |  |
| No risk | 29.5 | 260 | 10.5 | 118 | 14.9 | 221 | 14.9 | 371 | 16.2 | 971 |
| Nicotine abuse | 17.1 | 151 | 40.7 | 458 | 44.9 | 669 | 40.2 | 998 | 38.1 | 2,276 |
| Alcohol abuse | 4.0 | 35 | 3.1 | 35 | 2.5 | 37 | 1.4 | 35 | 2.4 | 142 |
| Immune suppression | 8.5 | 75 | 13.7 | 154 | 12.0 | 178 | 12.2 | 304 | 11.9 | 710 |
| Diabetes | 8.1 | 71 | 13.1 | 147 | 14.9 | 222 | 14.3 | 356 | 13.3 | 796 |
| Cancer | 5.6 | 49 | 11.7 | 131 | 9.1 | 135 | 10.3 | 255 | 9.5 | 570 |
| Pneumopathy | 2.3 | 20 | 2.8 | 31 | 2.4 | 35 | 0.4 | 11 | 1.6 | 97 |
| Nephropathy | 0.2 | 2 | 1.5 | 17 | 0.9 | 14 | 0.4 | 10 | 0.7 | 43 |
| Cardiopathy | 1.0 | 9 | 2.8 | 31 | 1.0 | 15 | 1.4 | 34 | 1.5 | 89 |
| Age 80+ years | 15.0 | 132 | 16.0 | 180 | 17.5 | 261 | 19.4 | 482 | 17.7 | 1,055 |
| **Diagnosis** |  |  |  |  |  |  |  |  |  |  |
| UAT | 86.4 | 761 | 83.2 | 935 | 85.6 | 1,274 | 82.0 | 2,036 | 83.8 | 5,010 |
| Culture | 7.3 | 64 | 6.0 | 67 | 4.7 | 70 | 4.6 | 114 | 5.3 | 314 |
| PCR | 3.3 | 29 | 7.4 | 83 | 8.6 | 128 | 10.6 | 263 | 8.4 | 503 |
| Serology | 4.7 | 41 | 2.9 | 33 | 2.2 | 32 | 0.7 | 17 | 2.1 | 123 |
| **Strains** |  |  |  |  |  |  |  |  |  |  |
| *L. pneumophila* | 95.5 | 840 | 95.2 | 1,072 | 96.0 | 1,429 | 95.4 | 2,371 | 95.5 | 5,712 |

LD: Legionnaires' disease; CI: Confidence interval; UAT: Urinary antigen test

Cases comprised of 68.9% (N=4,120) men, the mean age was 64 years (CI 95% 63-64). The age group of 60 to 69-year olds made up for one quarter of the legionellosis cases (22.7%). The notification rate of the whole period (2000-2020) was highest for the 80 to 89 years olds (13.3/100,000 population, Supplementary Table 1). The proportion of men among all cases was high over all years (range: 54.3% - 73.6%) and the overall and all period notification rates were more than double than those for women (5.0/100,000 versus 2.2/100’000 population). Over all study years, the canton of Ticino accounted for 15.0% of all cases, followed by the cantons of Zurich (14.0%) and Berne (10.2%). Yet, the notification rate in Ticino was found to be three to four times higher than the average of the other greater regions (Supplementary Table 1 and Supplementary Figure1). In 2020, fewer cases were reported from the cantons of Geneva (3.4%) and Neuchatel (1.5%) compared to their overall means (7.0% and 2.6%). In contrast, the canton of Valais reported more cases in 2020 (6.1%) than its overall mean (3.5%).

#### 3.2.2 Notification process

Of all cases, 91.9% (N=5,494) were classified as confirmed cases of LD, 1.3% (N=80) as probable and 6.8% (N=406) as possible cases. Congruently, 93.5% (N= 5,574) of all cases had both, a notification from the physician and from the diagnostic laboratory; 3.8% (N= 227) had only a laboratory notification and 0.1% (N= 4) were recorded with a clinical notification only. This proportion remained largely stable, however, in 2020, 8.1% (N= 39) of all cases were notified to the FOPH without a clinical notification form. This is in line with only 89.6% clinically confirmed LD cases in 2020, the lowest since 2000 (mean 2000-2020: 93.7%); and the highest number of cases classified as probable (11.2%, mean: 6.7%).

#### 3.2.3 Clinical information

Among all cases with a clinical notification form (N=5753), 85.8% were hospitalised in 2020 and in 2019 (mean: 89.9%). The median number of days from case date to hospitalisation was 3 days. The overall case fatality rate (CFR) was 5.2% (N=300). The annual CFR decreased from 7.7 % (CI: 2.5-17.0%) in 2000 to 3.6% (CI: 2.1 – 5.8%) in 2020. The CFR was highest in 2001 (10.2%, CI: 5.6 – 16.9%) and lowest in 2016 (2.8%, CI: 1.4 – 5.1%). The median duration from the reported case date to death was 7 days (10^th^ and 90^th^ percentile: 2-24 days). On average 97.4% of cases with a clinical report form were diagnosed with a pneumonia, thereby fulfilling the clinical criteria for diagnosing a LD.

#### 3.2.4 Exposure

If clinical reports were available, the risk factor for LD most often reported was nicotine abuse (39.6%), age being 80 and older (17.7%) and diabetes (13.8%). These proportions remained stable over the years after 2005. Most cases were classified as community-acquired (77.4.2%, N= 4,454) followed by travel-associated LD (13.8%, N=793), nosocomial (4.0%, N=231), related to a retirement home (2.9%, N=169) and occupation-related (1.8%, N=106). Among all travel-associated cases, the majority was traveling abroad (78.1%).

The proportion of travel-associated legionellosis cases most prominently decreased in 2020 (8.3%, mean: 13.8%), while the number of occupation-associated cases increased to 3.6% (mean: 1.8%). Further, the proportion of travels abroad decreased to 64.9% (mean: 78.1%).

#### 3.2.5 Diagnostics

Most cases were diagnosed using a urine sample (89.5%); sputum (6.4%), liquids from bronchoalveolar lavages (6.5%) and serum (2.2%) were significantly less often used. Consequently, the urinary antigen test (UAT) was used for most diagnostics (91.2%), followed by PCR (9.4%) and culture-based diagnostics (7.1%), and serological testing (3.2%). The proportion of PCR tests used increased continuously over the years. Of all 5,927 cases with the test specified, 642 (9.2%) had at least two different kinds of tests; the combinations of an UAT with a culture (N=315) and an UAT with a PCR test (N= 281) were most frequently recorded.

#### 3.2.6 *Legionella* species

Among all cases, *Legionella pneumophila* has been indicated as the causative agent for 95.5% (N=5,712). This proportion remained high across all years. If a culture or a PCR was indicated in the records, the species could be identified for 82.3% (730 out of 887). Of these, a significant proportion were identified as *Legionella pneumophila* (87.4%), among which serogroup 1 accounted for 21.5 %. Only 7 cases of *L. bozemanii*, 4 cases of *L. longbeachae*, 3 cases of *L. micdadei* and 1 case of *L. brunensis* infection were recorded.

### 3.3 Data quality of the NNSID database

#### 3.3.1 Completeness

The data between 2000 and 2020 was generally complete. In 2020, due to the reduced reporting of the clinical notification form, more clinical information was missing compared to previous years: the hospitalisation status was given only for 89.9% of all cases and the manifestation date (i.e. the date of disease onset) for 82.0%. A detailed overview is provided in Supplementary Table 2.

#### 3.3.2 Internal validity

Overall, the internal validity of the data was high and only a few inconsistencies were found. In 37 records (0.6%), the case classification and the entries of the clinical and laboratory criteria were discordant. From the cases with known disease onset date (N=5,111), 102 (2%) records indicated an onset date after the notification date. Similarly, in a few cases, the entries of date of death preceded the date of testing. We could not be evaluate the indicated exposure classification in relation to the incubation timeline.

#### 3.3.3 Timeliness

The median number of days between the case date to the hospitalisation date was 2 days (10 and 90 percentiles: 0-7 days). The median number of days between hospitalisation and reception of the notification at the FOPH was 5 days (10 and 90 percentiles: 1-16 days). On average, there was no delay between reception and data entry at the FOPH (0 days; 10 and 90 percentiles: 0-1 days).

In 2020, the median days between events has remained stable, however, the spread, i.e. the 90% percentile, increased, particularly during the peaks of the pandemic (spring and autumn 2020). The Supplementary Table 3 shows the overall median number of days from case date to notification entry at the FOPH.

### 3.4 Legionellosis notifications during 2020

The first cases of COVID-19 were identified in Switzerland in week 8 of 2020 (Figure 3a). The first wave of the pandemic peaked in week 12 with 7,118 cases and the second wave in week 44 with 56,093 cases. The most stringent non-pharmaceuticals measures (closure of schools, shops, sport centres and travel-restrictions) were set in place on March 16^th^ (week 13) and were then gradually removed until week 25. However, daily life was not resumed to levels before the pandemic between the first and second wave as some measures, such as quarantining if traveling from “risk countries” or limiting capacities at certain venues, persisted.

**Figure 3.**
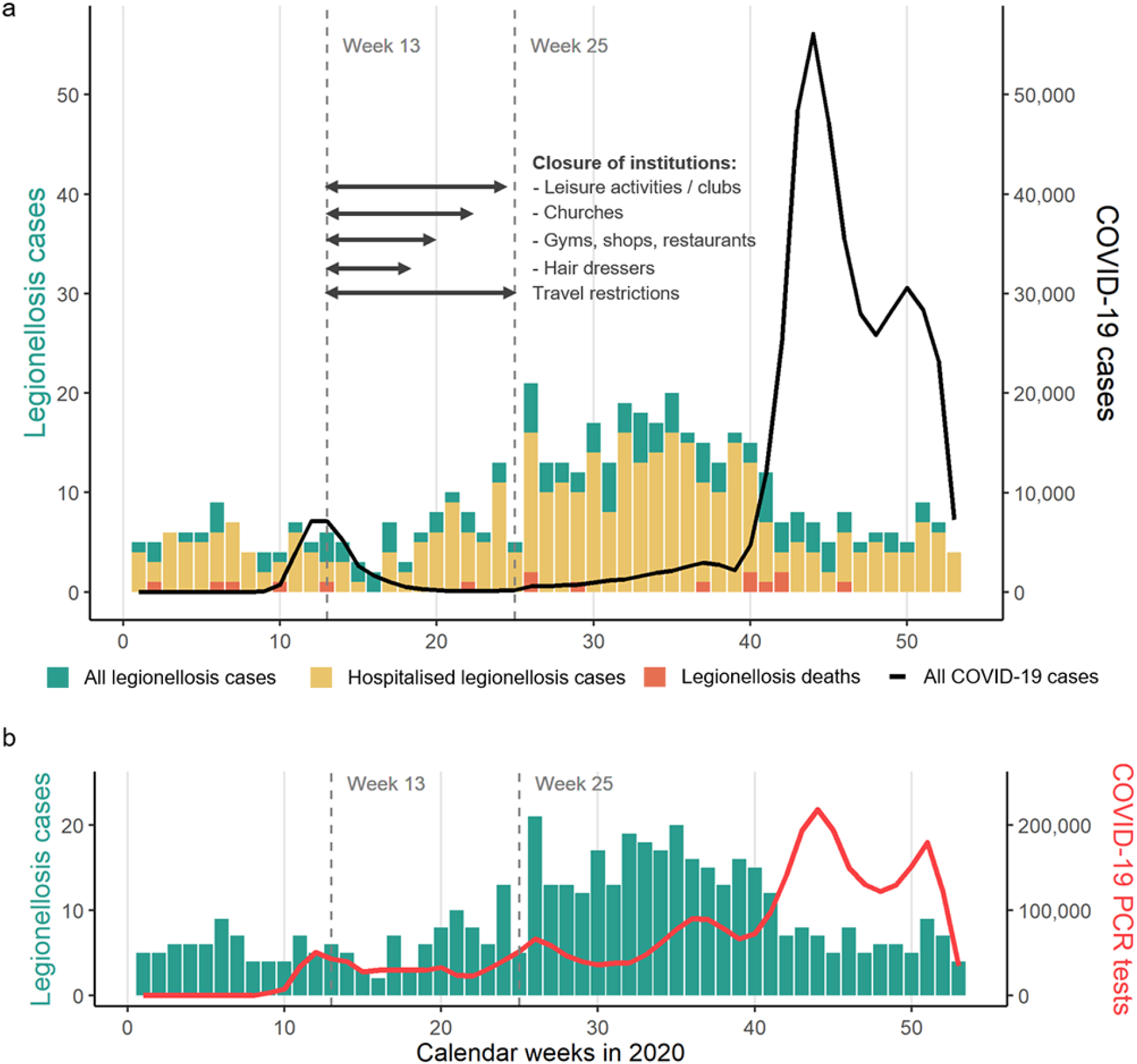
**a** Weekly number of legionellosis cases (left y-axis, scale 0-50) and COVID-19 cases (right y-axis, 0-50,000) in 2020, Switzerland. **b** Weekly number of legionellosis cases (left y-axis, scale 0-20) and COVID-19 PCR tests (right y-axis, scale 0-200,000) in 2020, Switzerland.

In total 483 legionellosis cases (among them 429 LD cases) were reported in 2020. In week 26 an early peak in legionellosis cases could be seen (21 cases), followed by the expected seasonal increase in cases by week 30/32. The number of legionellosis cases followed the usual seasonality with more cases occurring in summer than in winter. This contrasted with the period of relatively low COVID-19 incidence before the surge of the second wave.

Figure 3b illustrates the number of legionellosis cases and the frequency of COVID-19 PCR tests performed, which are weakly correlated (Spearman’s rank correlation= 0.38, p<0.01).

Figure 4 shows the results from the ITSA. The restriction of travel is associated with a decrease in notification rate of 35% (95% CI: 0.47-0.90; p< 0.01), the re-usage of buildings such as gyms, shops and restaurants (week 20) is statistically non-significantly associated with an 11% increase in notification rate (95% CI: 0.85-1.46; p= 0.44). Also all other opening steps were not associated with an increase in cases.

**Figure 4.**
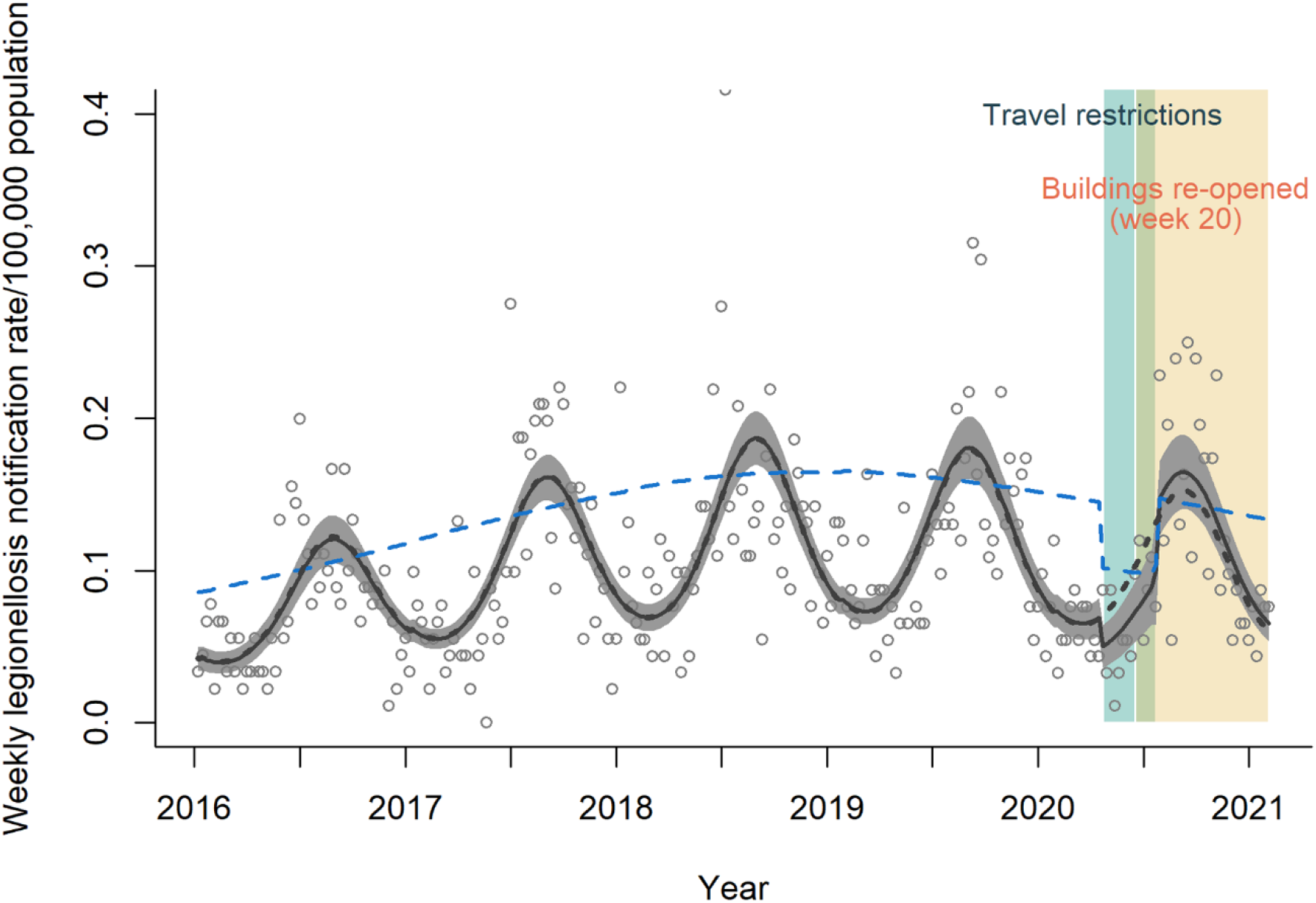
Interrupted time series analysis using Quasi-Poisson regression model on the number of weekly cases of legionellosis in Switzerland, 2016-2020. The blue line denotes the deseasonalised trend. The dotted black line represents the counterfactual if no interventions took place.

## 4 Discussion

### 4.1 Interruption of the upwards disease trend since 2018

We evaluated the Swiss legionellosis notification data over two decades. The upward trend since 2000 peaked in 2018 and plateaued thereafter. In 2018, the summer peak was also particularly strong and shifted into June instead of late summer time. This shift was most notably visible in Central Switzerland, Espace Mittelland, Northwestern Switzerland and to a lesser extent in the southern Swiss canton of Ticino. Therefore, this seasonal shift is unlikely driven by a cross-regional outbreak of legionellosis.

Comparing the most recent published European estimates on LD from 2019, Switzerland has one of the highest notification rates; Only Slovenia (9.4/100,000) reported higher rates [2]. While in about half of the European countries the upward trend in case notification after 2018 persisted, the strong and early summer peak in 2018 could be observed across all the EU/EEA and has been unmatched in 2019. The fact that also the US reported a similar high notification rate in 2018, suggest larger-scale (such as weather and climate) effects impacted LD occurrence [27]. The impact of climate, weather, relative humidity, and rainfall events in particular promoting LD infection and rising incidences has been highlighted before [28-30]. A study from the US attributed the peak in 2018 to climate changes but also increased road exposure (vehicle miles driven) [27]. Similarly, in Switzerland, spring (from April onward) and summer of 2018 were unusually warm and dry [31]. Yet, by the end of May several heavy rainfall and hail events occurred in the regions with the strongest summer peak [32].

The canton of Ticino exhibited a weekly notification rate three to four times higher compared to all other Swiss cantons. In addition, Ticino, out of all greater regions, experienced the strongest increase of LD since 2000 with pronounced surges in 2015 and 2018. A study investigating the number of diagnostic tests performed could show an increase in tests in the study period 2000-2017, yet also a high positivity rate suggesting that the notification numbers can not all be explained by testing behaviour [8]. The reason for the rise in incidence can only be hypothesised: During the summer months, Ticino had by far the most events of heavy rainfall [33]. Additionally, in the years 2015 and 2018, notably high levels of air pollution were registered in the Italian Po valley, affecting also the most southern valleys of Ticino [34].

It cannot be concluded yet, whether 2018 marks a turning point, if rates plateau or if 2018 is outlier in an otherwise continuing increasing trend in legionellosis notification rate. Data following the years after the extraordinary circumstances of the pandemic need to be closely monitored.

### 4.2 Stable risk groups and high level of data quality

There has been no remarkable shift in Swiss demographics across the years. The CFR for LD has been fluctuating throughout the years, but has been lower in recent years than at the start of the century. The overall CFR of 5% calculated from the NNSID data in our study is slightly lower than the average in the EU/EEA of 7% [2]. However, this figure needs to be interpreted with care: mandatory notification requires the information on the diagnostic (laboratory) findings and a report on clinical findings including exposure data and condition at time of reporting, but a follow-up reporting of the disease outcome including death is not mandatory. Given that notification often occurs early in the disease progression the LD-related CFR of 5% from NNSID data may be underestimated. Vital statistics are consistently collected at the FSO. The ICD-10 code A481 “Legionnaires’ disease” has been reported as primary or secondary cause of death for on average 23 cases per year (range 12 to 35 cases) in the decade from 2008 to 2018; (data provided by the FSO to the FOPH). Because the death reports do not always provide the underlying disease leading to respiratory or cardiovascular failure, they tend to underestimate the importance of infectious diseases as cause of death. Still, based on these estimates and for the reasons above the number of deaths in the NNSID was generally underestimated by on average of 30% (range 1%-58%).

Overall, the extent of data incongruities and missing data in the NNSID database is low, and notifications and data entry are made in a timely manner. Similar to the death status, other post-notification information on the development of the cases, such as the discharge date cannot be universally captured in the surveillance system. As a result, e.g. discharge date was removed from the reporting form in 2014. The median duration from requesting a diagnostic test for *Legionella* infection and legionellosis notification to the FOPH is 5 days and in due-time of the one week time limit for legionellosis notifications [35] and comparable to the Norwegian timeliness [36]. The variable “case date”, which fixes the case on the timeline does hamper the interpretation slightly as it can relate to various dates that were recorded within the disease progression. Finally, the current structure of the database is in part marginally user-friendly and/or has been changed (with little readily available documentation) over the years, impeding access to the information. For some reported information (e.g., exposure classification), the database does not allow automatic verification. Electronic reporting could support this process and facilitate data evaluation in the long term. Additionally, some of the incongruities might be avoidable if automated data checks would be included in such an electronic system at entry points with the laboratories and the physicians.

Lastly, the amount of information on each case has been decreasing in recent years with the omission of variables of the clinic progression and risk factors (e.g. occupation). Decreasing the requested information and streamlining the notification process to the data that is essential for the purpose of the surveillance, lowers the workload on the notifying physicians and might further improve (the already high) adherence and quality of the information provided.

### 4.3 The impact of COVID-19 on LD case numbers

In 2020, the first year of the SARS-CoV-2 pandemic, the number of reported legionellosis cases was similar to 2017. The epidemiological curves are nearly identical and lower than in 2018 and 2019 (see Supplementary Figure 2). A recent report from the FOPH noted a decline of LD cases of 32% compared to the expected case numbers based on the years 2015 to 2019 [9]. In our model, starting in 2016, the expected case numbers without the measures brought by the pandemic (the counterfactual) was lower than the actual case numbers. The interruption of the upwards trend in 2018 hampered accurate forward prediction and is dependent on the inclusion of years. However, the estimated effect of the investigated measures remained stable. The CFR was lower in 2020 than in any previous year, and a temporal pattern of reported deaths within 2020 could not be observed.

It is difficult to disentangle the effects of the pandemic on legionellosis notification rates. The pandemic itself had an influence on a multitude of aspects of our life and the main causes of LD are not well understood yet. There could be factors that have generally not been considered, e.g. the reduction of air pollution [37], which may well be part of the causal pathway for LD. For example, it has been previously hypothesised that high concentrations of PM_10_ is associated with an increase of viable bacteria in the air [38] and increased LD incidence [39, 40].

A notable difference in 2020, however, was a 4-percentage-point reduction of clinical notification forms submitted to the NNSID. Therefore, less information to evaluate on hospitalisation and other clinical attributes were available in 2020. The clinical notification is sent by the treating physician to the cantonal physicians, who processes and forwards the notification to the FOPH [5]. In case, the cantonal physician receives a laboratory but no clinical notification, he requests a clinical notification from the treating physician. These clinical notification forms are most prominently missing in April and October 2020, suggesting (hospital) physicians and/or cantonal authorities were preoccupied with the consequences and the control of the COVID-19 pandemic while the notification through the diagnostic laboratories was able to continue. The Swiss notification system records “possible” cases as legionellosis, but not as LD. This leads to an underestimation of LD cases, if the clinical report is missing or, if information on the disease progression and thereby the occurrence of pneumonia is incomplete at time of reporting.

There has been a notable decrease in travel-associated cases during the pandemic, associated with both, domestic and international travel [11]. Concurrently, the ITSA showed a marked drop in legionellosis cases associated with the implementation of travel restrictions; and after these were lifted, there was a corresponding increase in cases. The travel restrictions also seemed to have a stronger effect on the case numbers than the re-opening of buildings and the suspected subsequent exposure of the population to higher *Legionella* spp. concentrations. According to a recent article water stagnation related issues following closure of buildings might have overestimated respective overstated the risk for LD, as the proliferation of *Legionella* spp. is dependent on several other factors (e.g. nutrients, temperature) [41]. Yet, there is no concluding evidence for either side. The lack of effect could also be due to staggered re-opening of buildings, spreading the new cases and diluting the effect, or flushing recommendations in anticipation of the risk through stagnation have been taken seriously by buildings owners / management and cases were successfully prevented.

We found a weak correlation between the number of COVID-19 tests performed and the number of LD cases identified. Due to the similar clinical representations, this was expected. In Switzerland, patients presenting at the emergency ward or admitted to the hospital with severe pneumonia, are recommended to be tested for *Legionella* spp. using the UAT [42]. Co-infections of COVID-19 and LD has also been described in various studies (1-3%) although the sample size was often rather small [43, 44].

### 4.4 Conclusion

In Switzerland, the notification rate of LD continuously increased since 2000 to one of the highest rates in Europe, yet the upwards trends was interrupted in 2018, the reason remains unclear. The COVID-19 pandemic seemed to have affected the case numbers mainly through the travel restrictions, which has notably decreased the number of travel-associated cases. Additionally, while physicians seemed to lack resources to keep up with their obligations to notify, the notifications were reported through the diagnostic laboratories in similar frequency and quality compared to previous years, suggesting a robust surveillance system.

### 4.5 Limitations

As this study was based on information from passive disease surveillance, we were limited to cases that were reported. Therefore, we could only approximate the true incidence of the disease. Further, the main drawback on studies involving surveillance data is the lack of denominator data. However, a study on this additional data for the years 2007-2016 has been published previously [8].

## Data Availability

All data are available upon reasonable request to the authors and upon agreement of the Federal Office of Public Health.

## Acknowledgements

We thank Dr. Jan Hattendorf (Swiss TPH), Anja Orschulko (Swiss TPH), Julia Fanderl (Swiss TPH) and Dr. Monica Golumbeanu (Swiss TPH) for their support in the statistical analysis, data acquisition, data cleaning and advice. We also thank the Federal Office of Public Health, in particular Marianne Jost, Ornella Luminati and Dr. Ekkehardt Altpeter for providing the data and their support.

## 6 Statements and Declarations

### 6.1 Funding

This study was funded by the Federal Office of Public Health (FOPH, contract number 142003961 / 334.0-85/53).

### 6.2 Competing Interests

The authors have no conflicts of interest to declare that are relevant to the content of this article. Monica N. Wymann is staff of the FOPH and participated in her capacities as public health specialist and her function as scientific collaborator within the organisation.

### 6.3 Author contributions

All authors contributed to the study conception and design. Material preparation and data collection were performed by Fabienne B. Fischer and Monica N. Wymann. Analysis and interpretation was performed by Fabienne B. Fischer with support of Daniel Mäusezahl and Monica N. Wymann. The first draft of the manuscript was written by Fabienne B. Fischer and all authors commented on previous versions of the manuscript. All authors read and approved the final manuscript.

### 6.4 Ethics approval

The study was conducted under the Epidemics Act (SR 818.101) [35]. The study team received the legionellosis notification data from the FOPH. Other data (COVID-19 cases, non-pharmaceutical measures, and population statistics) are publicly available from the FOPH, the FSO or third parties.

### 6.5 Data availability

All data are available upon reasonable request to the authors and upon agreement of the FOPH.

## Footnotes

1 The annual crude notification rate for LD cases (legionellosis cases with confirmed pneumonia) ranged from 0.9/100,000 population (CI: 0.7 - 1.1) in 2000 to 4.9/100,000 population (CI: 4.5 – 5.4) in 2020. The highest notification rate was recorded in 2018 with 6.3/100,000 population (CI: 5.8 – 6.9).

## Notes

### Funding Statement

This study was funded by the Federal Office of Public Health (contract number 142003961 / 334.0-85/53).

### Author Declarations

The study was conducted under the Epidemics Act (SR 818.101) [35]. The study team received the legionellosis notification data from the Federal Office of Public Health (FOPH). Other data (COVID-19 cases, non-pharmaceutical measures, and population statistics) are publicly available from the FOPH, the Federal Statistical Office or third parties.

### Summary of Updates

The authors have been re-ordered in the system to: Fabienne B. Fischer, Daniel Maeusezahl and Monica N. Wymann. The author order in the manuscript was correct. The uploaded manuscript is the same as before, but we had to re-upload in order to complete the submission form.

## References

1. Thacker SB, Choi K, Brachman PS. The surveillance of infectious diseases. JAMA. 1983;249(9):1181–5.

2. European Centre for Disease Prevention and Control. Legionnaires’ disease. In: ECDC. Annual epidemiological report for 2019. Stockholm: ECDC; 2021.

3. Centers for Disease Control and Prevention. Legionnaires’ disease surveillance summary report, United States: 2016 - 2017. US: CDC; 2020.

4. Fukushima S, Hagiya H, Otsuka Y, Koyama T, Otsuka F. Trends in the incidence and mortality of legionellosis in Japan: a nationwide observational study, 1999–2017. Sci. Rep. 2021;11(1):7246. doi:10.1038/s41598-021-86431-8

5. Schmutz C. Foodborne diseases in Switzerland: Understanding the burden of illness pyramid to improve Swiss infectious disease surveillance [Doctoral dissertation]. Swiss Tropical and Public Health Institute and University of Basel, Basel, Switzerland; 2018.

6. Bundesamt für Gesundheit. Meldepflichtige übertragbare Krankheiten und Erreger: Leitfaden zur Meldepflicht. 2020.

7. Gysin N. Legionnaires’ disease in Switzerland: analysis of Swiss surveillance data, 2000 to 2016 - spatial and seasonal determinants [MPH thesis]. 2018.

8. Fischer FB, Schmutz C, Gaia V, Mäusezahl D. Legionnaires’ disease on the rise in Switzerland: A denominator-based analysis of national diagnostic data, 2007–2016. Int. J. Environ. Res. Public Health. 2020;17(19):7343.

9. Der Einfluss der durch COVID-19-bedingten Massnahmen und Verhaltensänderungen auf meldepflichtige Infektionskrankheiten in der Schweiz im Jahr 2020. BAG Bulletin 30/2021: Bundesamt für Gesundheit; 2021.

10. Cassell K, Davis JL, Berkelman R. Legionnaires’ disease in the time of COVID-19. Pneumonia. 2021;13(1):2. doi:10.1186/s41479-020-00080-5

11. Steffen R, Lautenschlager S, Fehr J. Travel restrictions and lockdown during the COVID-19 pandemic—impact on notified infectious diseases in Switzerland. J. Travel Med. 2020. doi:10.1093/jtm/taaa180

12. Palazzolo C, Maffongelli G, D’Abramo A, et al. Legionella pneumonia: increased risk after COVID-19 lockdown? Italy, May to June 2020. Euro Surveill. 2020;25(30):2001372. doi: 10.2807/1560-7917.ES.2020.25.30.2001372

13. Proctor CR, Rhoads WJ, Keane T, et al. Considerations for large building water quality after extended stagnation. AWWA Water Sci. 2020;2(4):e1186. doi:10.1002/aws2.1186

14. ESCMID Study Group for Legionella Infections. ESGLI Guidance for managing Legionella in building water systems during the COVID-19 pandemic. 2020.

15. Dey R, Ashbolt NJ. Legionella infection during and after the COVID-19 pandemic. ACS ES&T Water. 2020. doi:10.1021/acsestwater.0c00151

16. FDHA Ordinance of 16 December 2016 on Drinking Water and Water in Public Baths and Shower Facilities (SR 817.022.11). 2017.

17. Federal Office of Public Health. Legionellose. In: Zahlen zu Infektionskrankheiten. 2021. https://www.bag.admin.ch/bag/de/home/zahlen-und-statistiken/zahlen-zu-infektionskrankheiten.exturl.html/aHR0cHM6Ly9tZWxkZXN5c3RlbWUuYmFnYXBwcy5jaC9pbmZyZX/BvcnRpbmcvZGF0ZW5kZXRhaWxzL2QvbGVnaW9uZWxsYS5odG1s/P3dlYmdyYWI9aWdub3Jl.html. Accessed 4 May 2021.

18. Legionellen und Legionellose BAG-/BLV-Empfehlungen: Federal Office of Public Health; Federal Food Safety and Veterinary Office. 2018.

19. Hale T, Angrist N, Goldszmidt R, et al. A global panel database of pandemic policies (Oxford COVID-19 Government Response Tracker). Nat. Hum. Behav. 2021.

20. COVID_measures_CH. https://github.com/SwissTPH/COVID_measures_by_canton. Accessed 13 February 2021.

21. Federal Authorities. COVID-19 Switzerland. 2021. https://www.covid19.admin.ch/en/overview. Accessed 18 March 2021.

22. R Core Team. R: A Language and Environment for Statistical Computing. Vienna, Austria: R Foundation for Statistical Computing; 2020.

23. Fay MP, Feuer EJ. Confidence intervals for directly standardized rates: a method based on the gamma distribution. Stat. Med. 1997;16(7):791–801.

24. Bernal JL, Cummins S, Gasparrini A. Interrupted time series regression for the evaluation of public health interventions: a tutorial. Int. J. Epidemiol. 2017;46(1):348–55. doi:10.1093/ije/dyw098

25. Ordinance 2 of 13 March 2020 on Measures to Combat the Coronavirus (SR 818.101.24). 2020.

26. Berkelman R. Legionellosis. In: Control of Communicable Diseases Manual. 2015.

27. Han XY. Effects of climate changes and road exposure on the rapidly rising legionellosis incidence rates in the United States. PLoS One. 2021;16(4):e0250364. doi:10.1371/journal.pone.0250364

28. Sakamoto R. Legionnaire’s disease, weather and climate. Bull. World Health Organ. 2015;93(6):435–6. doi:10.2471/BLT.14.142299

29. Gleason JA, Kratz NR, Greeley RD, Fagliano JA. Under the weather: legionellosis and meteorological factors. Ecohealth. 2016;13(2):293–302. doi:10.1007/s10393-016-1115-y

30. Walker JT. The influence of climate change on waterborne disease and Legionella: a review. Perspect Public Health. 2018;138(5):282–6. doi:10.1177/1757913918791198

31. MeteoSchweiz. Klimabulletin Jahr 2018. Zürich: Bundesamt für Meteorologie und Klimatologie. 2019.

32. MeteoSchweiz. Klimareport 2018. Zürich: Bundesamt für Meteorologie und Klimatologie. 2019.

33. MeteoSchweiz. Starkniederschläge. Bundesamt für Meteorologie und Klimatologie. 2018. https://www.meteoschweiz.admin.ch/home/klima/schweizer-klima-im-detail/starkniederschlaege.html. Accessed 16 June 2021.

34. European Environment Agency. Air quality in Europe - 2020 report. EEA Report No 9/2020. 2020.

35. Communicable Diseases Legislation – Epidemics Act (SR 818.101). 2016.

36. Wolff C, Lange H, Feruglio S, Vold L, MacDonald E. Evaluation of the national surveillance of Legionnaires’ disease in Norway, 2008-2017. BMC Public Health. 2019;19(1):1624. doi:10.1186/s12889-019-7981-9

37. Venter ZS, Aunan K, Chowdhury S, Lelieveld J. COVID-19 lockdowns cause global air pollution declines. Proc. Natl. Acad. Sci. U. S. A. 2020;117(32):18984. doi:10.1073/pnas.2006853117

38. Mancinelli RL, Shulls WA. Airborne bacteria in an urban environment. Appl. Environ. Microbiol. 1978;35(6):1095–101.

39. Halsby K, Joseph C, Lee J, Wilkinson P. The relationship between meteorological variables and sporadic cases of Legionnaires’ disease in residents of England and Wales. Epidemiol Infect. 2014;142(11):2352–9.

40. Russo A, Gouveia CM, Soares PM, Cardoso RM, Mendes MT, Trigo RM. The unprecedented 2014 Legionnaires’ disease outbreak in Portugal: atmospheric driving mechanisms. Int. J. Biometeorol. 2018;62(7):1167–79.

41. Rhoads WJ, Hammes F. Growth of Legionella during COVID-19 lockdown stagnation. Environ Sci: Water Res Technol. 2021;7(1):10–5. doi:10.1039/D0EW00819B

42. Ewig S, Kolditz M, Pletz M, et al. Behandlung von erwachsenen Patienten mit ambulant erworbener Pneumonie–Update 2021. Pneumologie. 2021;75(09):665–729.

43. Verhasselt HL, Buer J, Dedy J, et al. COVID-19 Co-infection with Legionella pneumophila in 2 Tertiary-Care Hospitals, Germany. Emerg. Infect. Dis. 2021;27(5):1535.

44. Adler H, Ball R, Fisher M, Mortimer K, Vardhan MS. Low rate of bacterial co-infection in patients with COVID-19. The Lancet Microbe. 2020;1(2):e62. doi:10.1016/S2666-5247(20)30036-7.

