## Supplementary file containing Supplementary Tables 1-3 and Supplementary Figures 1 and 2. for "Temporal trends in legionellosis national notification data and the effect of COVID-19, Switzerland, 2000- 2020" (from another version of this preprint: version 1, 2022-01-21)

### Supplementary information

**Supplementary Table 1** Period notification rates and 20-year notification rate for legionellosis in Switzerland, 2000-2020

| | Notification rate per 100,000 population for each period | | | | |
|---|---|---|---|---|---|
| | 2000-2005 | 2006-2010 | 2011-2015 | 2016-2020 | Overall |
| ***Age category (years)*** | | | | | |
| 0 to 19 | 0.1 | 0.1 | 0.1 | 0.1 | 0.1 |
| 20 to 29 | 0.2 | 0.6 | 0.4 | 0.9 | 0.5 |
| 30 to 39 | 0.8 | 1.0 | 1.2 | 1.9 | 1.2 |
| 40 to 49 | 1.9 | 2.4 | 2.7 | 3.7 | 2.6 |
| 50 to 59 | 3.0 | 4.3 | 5.5 | 8.0 | 5.3 |
| 60 to 69 | 4.1 | 6.3 | 7.9 | 12.3 | 7.8 |
| 70 to 79 | 6.4 | 8.6 | 9.7 | 14.9 | 10.1 |
| 80 to 89 | 7.3 | 10.0 | 12.7 | 21.9 | 13.3 |
| 90+ | 4.8 | 8.3 | 14.6 | 21.1 | 12.8 |
| ***Sex*** | | | | | |
| Male | 2.8 | 4.2 | 5.1 | 8.0 | 5.0 |
| Female | 1.3 | 1.7 | 2.2 | 3.7 | 2.2 |
| ***Region*** | | | | | |
| Central Switzerland | 0.0 | 0.1 | 0.1 | 0.3 | 0.1 |
| Eastern Switzerland | 0.1 | 0.1 | 0.2 | 0.2 | 0.1 |
| Espace Mittelland | 0.1 | 0.2 | 0.2 | 0.3 | 0.2 |
| Lake Geneva | 0.1 | 0.2 | 0.2 | 0.3 | 0.2 |
| Northwestern Switzerland | 0.1 | 0.2 | 0.2 | 0.3 | 0.2 |
| Ticino | 0.4 | 0.6 | 0.7 | 1.2 | 0.7 |
| Zurich | 0.1 | 0.1 | 0.1 | 0.3 | 0.2 |

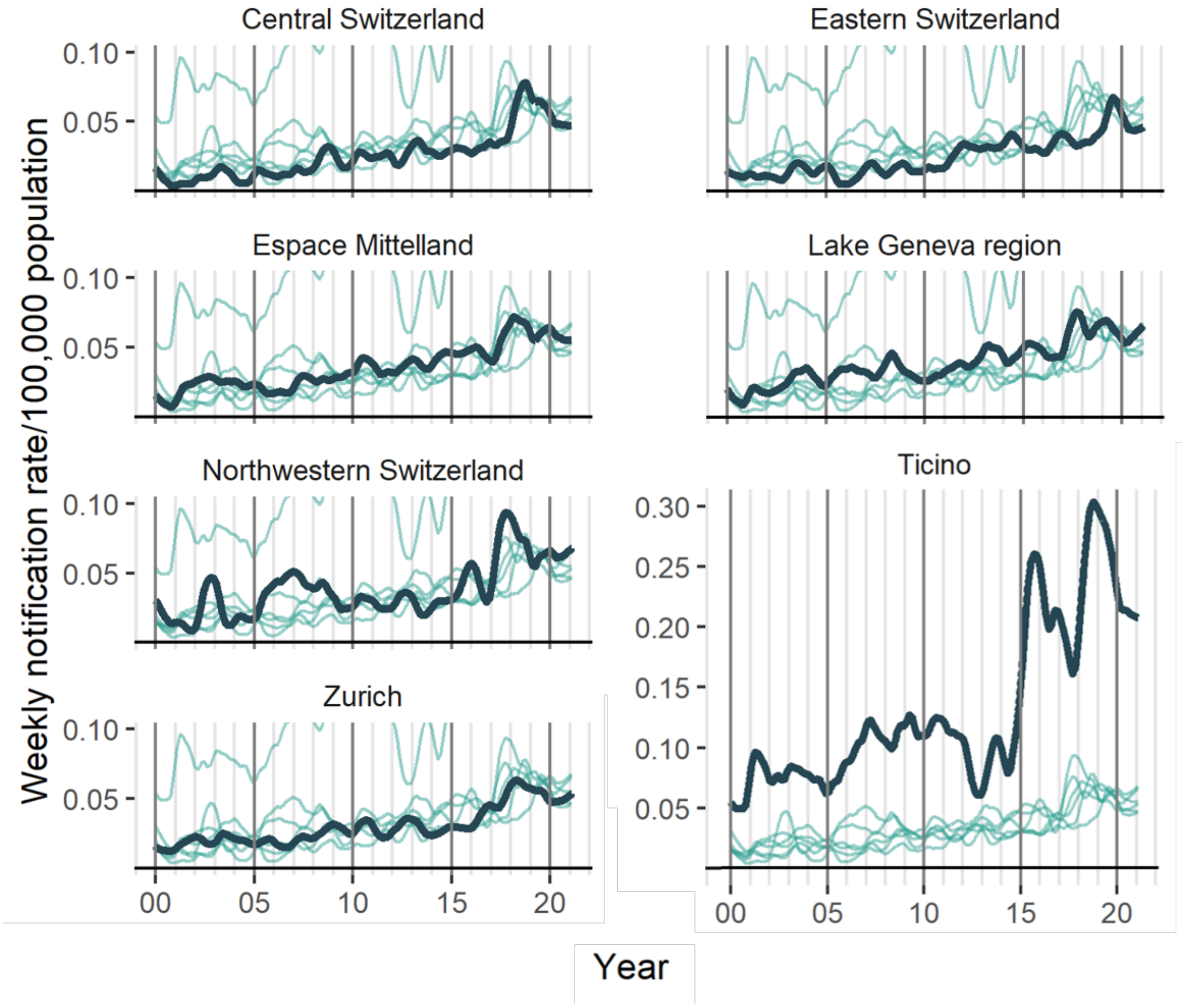


**Supplementary Figure 1** Weekly notification rate for legionellosis between 2000 and 2020 across the seven greater regions (NUTS-2 level) in Switzerland. The think green line denotes the notification rate of the specific region, while the thinner lines are the notification rate of all other six regions for easier visual comparison.

**Supplementary Table 2** Completeness of key variables for the notification of legionellosis in Switzerland with 95% confidence interval across all years, for all cases in the database and confirmed and probable cases only, 2000-2020

| | **All cases (N=5,980)** | | | | **Confirmed and probable cases (N=5,578)** | | | |
|---|---|---|---|---|---|---|---|---|
| | Variable not empty | | Variable not empty and not "unknown" | | Variable not empty | | Variable not empty and not "unknown" | |
| | % | *95% CI* | % | *95% CI* | % | *95% CI* | % | *95% CI* |
| ***Notification*** | | | | | | | | |
| Case classification | 100.0 | *100-100* | - | - | 100.0 | *100-100* | - | - |
| Clinical information | 100.0 | *100-100* | - | - | 100.0 | *100-100* | - | - |
| Laboratory information | 100.0 | *100-100* | - | - | 100.0 | *100-100* | - | - |
| Name of reporting institution | 81.7 | *74.3-89.2* | 65.2 | *53.5-76.8* | 86.2 | *78.0-94.5* | 68.2 | *56-80.4* |
| ***Demographics*** | | | | | | | | |
| Age | 99.93 | *99.85-100* | - | - | 99.98 | *99.94-100* | - | - |
| Sex | 99.98 | *99.94-100* | - | - | 99.97 | *99.94-100* | - | - |
| Canton of residence | 99.6 | *99.4-99.7* | - | - | 99.7 | *99.5-99.9* | - | - |
| Nationality | 82.6 | *80.0-85.1* | 82.3 | *79.8-84.9* | 86.7 | *84.8-88.7* | 86.4 | *84.44-88.45* |
| ***Clinic*** | | | | | | | | |
| Hospitalisation | 100 | *100-100* | 91.6 | *88.9-94.4* | 100.0 | *100-100* | 96.4 | *94.8-98.0* |
| Hospitalisation date | 98.4 | *97.8-98.9* | - | - | 98.4 | *97.9-99* | - | - |
| Discharge date* | 61.7 | *57.1-66.3* | - | - | 61.5 | *56.8-66.1* | - | - |
| Death | 100.0 | *100-100* | - | - | 100.0 | *100-100* | - | - |
| Date of death | 81.9 | *77.4-86.4* | - | - | 82.5 | *78-87.1* | - | - |
| Manifestation date | 84.7 | *82.3-87.1* | - | - | 89.5 | *88.2-90.8* | - | - |
| Pneumonia | 94.9 | *93.1-96.6* | - | - | 99.95 | *99.9-100* | - | - |

| | | | | | | | | |
|---|---|---|---|---|---|---|---|---|
| ***Exposure*** | | | | | | | | |
| Risk factor | 100.0 | *100-100* | 84.7 | *82.4-86.9* | 100.0 | *100-100* | 88.6 | *86.6-90.5* |
| Modus | 100.0 | *100-100* | - | - | 100.0 | *100-100* | - | - |
| National or international travel if travel-associated LD | 100.0 | *100-100* | 92.0 | *88.6-95.4* | 100.0 | *100-100* | 92.9 | *89.6-96.3* |
| ***Diagnostic*** | | | | | | | | |
| Diagnostic test | 90.9 | *89.4-92.5* | 90.9 | *89.4-92.5* | 92.9 | *91.5-94.3* | 92.9 | *91.5-94.3* |
| Sample material | 93.8 | *92.6-95.0* | 93.8 | *92.6-95.0* | 93.6 | *92.3-94.9* | 93.6 | *92.3-94.9* |

* Variable omitted from the notification form in 2014

**Supplementary Table 3** Number of days between different steps to notification for legionellosis in Switzerland, 2000-2020

| | | 2000-2005 | | 2006-2010 | | 2011-2015 | | 2016-2020 | | Overall | |
|---|---|---|---|---|---|---|---|---|---|---|---|
| | N | p50[a] | (p10, p90)[b] | p50 | (p10, p90) | p50 | (p10, p90) | p50 | (p10, p90) | p50 | (p10, p90) |
| Case date to diagnostic sampling | 3,782 | 0 | (0, 0) | 3 | (0, 8) | 3 | (0, 8) | 3 | (0, 7) | 3 | (0, 7.9) |
| Diagnostic sampling to notification | 3,782 | 3 | (-1, 8.4) | 7 | (2, 20) | 5 | (1, 13) | 4 | (1,11) | 5 | (1, 13) |
| Reception of notification to data entry | 5,976 | 0 | (0, 0) | 0 | (0, 1) | 0 | (0, 0) | 0 | (0, 2) | 0 | (0, 1) |
| Case date to data entry | 5,976 | 7 | (0, 22) | 10 | (4, 26) | 8 | (3, 20) | 8 | (3, 19) | 8 | (2, 20) |

[a] 50th percentile / median

[b] 10th and 90th percentile

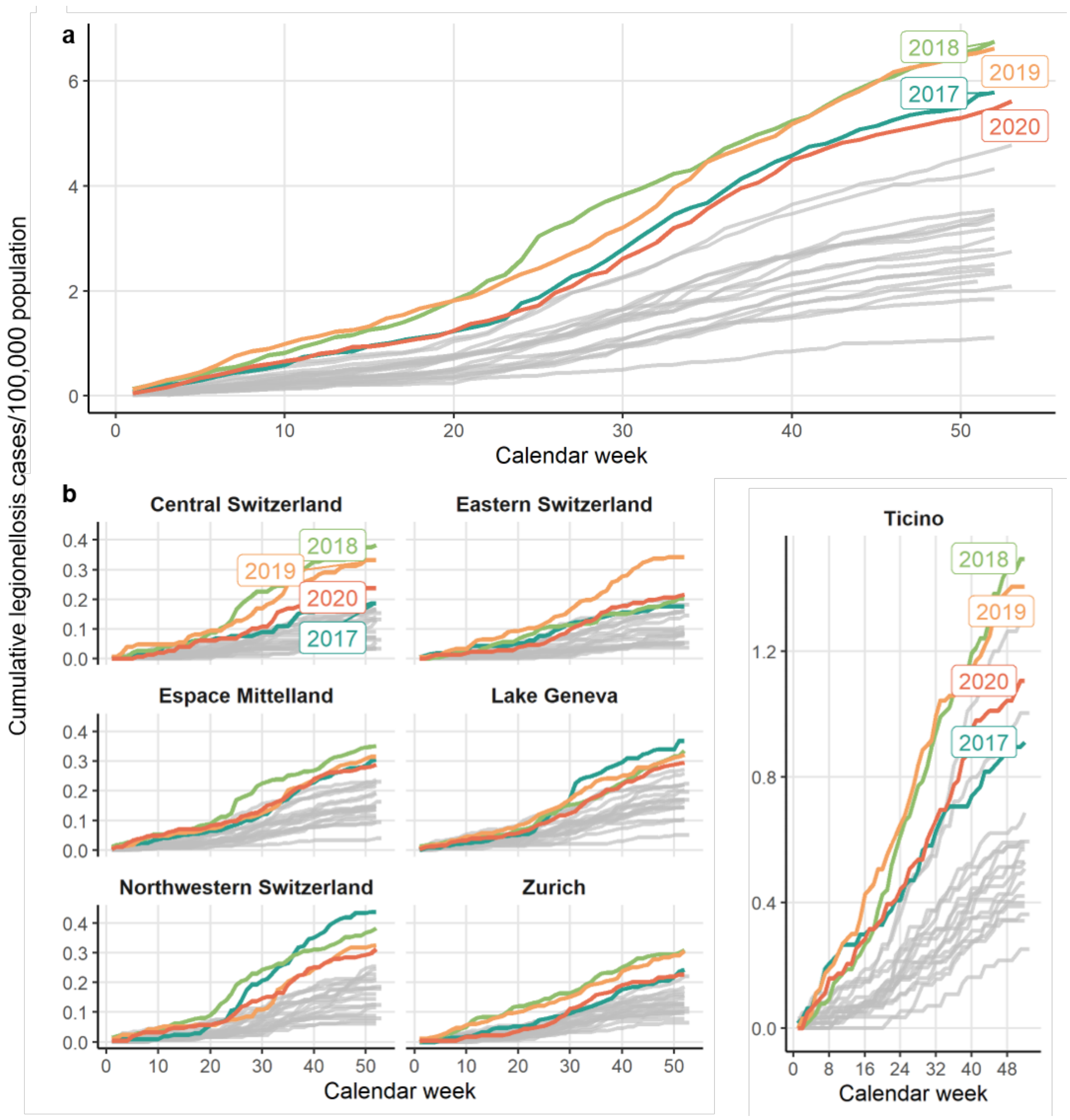


**Supplementary Figure 2** Cumulative legionellosis case number across the calendar year since 2000 across Switzerland (a) and stratified by greater region (NUTS-2 level, b). Highlighted are the last 4 years. The reference population for the notification rate is constant over the calendar year.
